# A phase 3, randomised, double-blind, placebo-controlled clinical trial for adult evaluation of the efficacy and safety of a SARS-CoV-2 recombinant spike RBD protein vaccine (ABDALA-3 Study)

**DOI:** 10.1101/2022.09.08.22279690

**Authors:** Francisco Hernández-Bernal, Maria C Ricardo-Cobas, Yenima Martín-Bauta, Ernesto Rodríguez-Martínez, Klaudia Urrutia-Pérez, Karen Urrutia-Pérez, Joel Quintana-Guerra, Zadis Navarro-Rodríguez, Marjoris Piñera-Martínez, José L Rodríguez-Reinoso, Cristina O Chávez-Chong, Idania Baladrón-Castrillo, Grettel Melo-Suárez, Alejandro Batista-Izquierdo, Alexis Pupo-Micó, Ricardo Mora-Betancourt, Dayamí Soler-Cano, Jacqueline Bizet-Almeida, Maria C Martínez-Rodríguez, Leonardo Lobaina-Lambert, Vivian M Velázquez-Pérez, Jalimy Soler-Díaz, Yunaili Blanco-Garrido, Sandra Laurencio-Vallina, Tamara Meriño-Hechavarría, Norberto Carmenaty-Campos, Enri Rodríguez-Montero, Miladys Limonta-Fernández, Marel Alonso-Valdés, Reinier Hernández-Rodríguez, Eulogio Pimentel-Vázquez, Karem M Catasús-Álvarez, Maria V Cabrera-Núñez, Marta Ayala-Ávila, Verena L Muzio-González, for the ABDALA Group of Investigators

**Author notes:** **Corresponding author:** Prof Francisco Hernández-Bernal, Clinical Research Direction, Centre for Genetic Engineering and Biotechnology (CIGB), P.O. Box 6162, Havana, Cuba. Contributed equally (Francisco Hernández-Bernal, Verena L Muzio-González). Members of the study group are listed in the Appendix.

## Abstract

**Background:** The pandemic of COVID-19 raised the urgent need of safe and efficacious vaccines against SARS-CoV-2. We evaluated the efficacy and safety of a new SARS-CoV-2 virus receptor-binding domain (RBD) vaccine.

**Methods:** A phase 3, multicentre, randomised, double-blind, placebo-controlled trial was carried out at 18 clinical sites in three provinces of the south-eastern region of Cuba. Subjects (healthy or those with controlled chronic diseases) aged between 19 and 80 years, who gave written informed consent were eligible. Subjects were randomly assigned (1:1, in blocks) to two groups: placebo, and 50 µg RBD vaccine (Abdala). The product was administered intramuscularly, 0.5 mL in the deltoid region, in a three dose immunization schedule at 0-14-28 days. The organoleptic characteristics and presentations of vaccine and placebo were identical. All participants (subjects, clinical researchers, statisticians, laboratory technicians, and monitors) remained blinded during the study period. The main endpoint was to evaluate the efficacy of the Abdala vaccine in the prevention of symptomatic COVID-19. The trial is registered with the Cuban Public Registry of Clinical Trials, RPCEC00000359.

**Findings:** Between March 22 to April 03, 2021, 48290 subjects were included (24144 and 21146 in the placebo and Abdala groups, respectively). The product was well tolerated. No severe adverse events with demonstrated cause-effect relationship attributable to vaccine were reported. The incidence of adverse reactions in the placebo and Abdala vaccine arms were 446/24144 (1.9%) and 615/24146 (2.5%), respectively. Adverse reactions were mostly mild, and from the injection site, which resolved in the first 24-48 hours. The Abdala vaccine efficacy against symptomatic COVID-19 was 92.28% (95% CI 85.74-95.82). In the case of mild/moderate disease the vaccine efficacy was 91.96% (84.69-95.78) and 94.46% (58.52-99.28) for the severe forms (serious/critical disease). There were five critical patients (of which four died), all in the placebo group, indicating that Abdala vaccine efficacy for both conditions was of 100%.

**Interpretation:** The Abdala vaccine was safe, well tolerated, and highly effective, fulfilling the WHO target product profile for COVID-19 vaccines.

**Funding:** Centre for Genetic Engineering and Biotechnology (CIGB), Havana, Cuba.

## Introduction

The COVID-19 pandemic is caused by severe acute respiratory syndrome coronavirus 2 (SARS-CoV-2). As of 21 August 2022, 593 million confirmed cases and 6·4 million deaths have been reported globally, according to data recorded by the World Health Organization (WHO).^1^

There is consensus that, as long as no safe and effective preventive vaccines are available for SARS-CoV-2, in sufficient quantities to implement comprehensive immunization programmes, the world will not return to normality. Vaccines are urgently needed to mitigate the consequences of this pandemic and protect humanity from future epidemics caused by this virus.^2^

Abdala vaccine was designed at CIGB, considering the state-of-the-art of research around COVID-19, especially the immunological aspects necessary for the development of vaccines against this infection. This vaccine is based on the recombinant RBD subunit of the spike protein produced in *Pichia pastoris* yeast, adjuvanted to alumina^3,4^ which is a very well-known platform previously used by this institution.

An exploratory phase 1-2 study, carried out in adults 19-80 years of age, evaluated the safety and immunogenicity of the Abdala vaccine. The vaccine was safe, well-tolerated and induced strong humoral immune responses against SARS-CoV-2.^5^ The aim of the present work was to evaluate the efficacy and safety of the Abdala vaccine, in the prevention of symptomatic disease due to SARS-CoV-2 infection in adults 19-80 years of age.

## Methods

### Study design

A phase 3, multicentre, randomised, double-blind, placebo-controlled clinical trial was carried out at 18 clinical sites in three provinces of the south-eastern region of Cuba.

The trial was conducted in medical wards and certified areas for the vaccination process. The clinical investigators were specialists in family medicine, internal medicine, and critical care, and the study vaccination was administered by specialist nurses. The protocol followed the guidelines of the Declaration of Helsinki and was evaluated by an *ad hoc* Centralized Ethics Committee, integrated by members of the Ethics and Review Committee of the Provincial Hospital “Saturnino Lora” in Santiago de Cuba (main clinical site), extended with members of the Research Ethics Committees of the Universities of Medical Sciences of Santiago de Cuba, Guantánamo and Granma, who granted ethical approval of the study. This Review Boards was made up of highly qualified medical specialists not linked to the study, as well as a member of the community. This committee followed up on the research ensuring the protection of the rights, safety and well-being of the subjects involved in the study. In addition, the Cuban Centre for State Control of Drugs, Medical Devices and Equipment approved the start of the clinical trial after considering the scientific, methodological and ethical aspects. The manuscript adheres to CONSORT reporting guidelines.

### Participants

Subjects aged between 19 and 80 years (healthy adults or with comorbidities, compensated), who gave their written, informed consent to participate, were enrolled. Key exclusion criteria were: subjects with previous infection to SARS-CoV-2 confirmed by reverse-transcriptase polymerase chain reaction (RT-PCR) or at the moment of inclusion, individuals suspicious to have the infection or in contact with a COVID-19 case, or previous any acute infections in the last 15 days, chronic diseases decompensated at the time of inclusion, subject who had received a vaccine candidate against COVID-19 or with any medical condition that requires an immunomodulator, systemic steroid or cytostatic during the study. Full details regarding the inclusion and exclusion criteria are provided in the clinical trial protocol (Appendix).

### Randomisation and masking

The subjects included were randomly distributed (1:1) to 2 groups: I) placebo and II) 50 µg RBD (Abdala vaccine). Randomisation was carried out in the supply group of the Clinical Research Direction of CIGB, in blocks of 4 individuals, for each clinical site, by means of a computerised random number generator. The sites received the product in such blocks, in masked vials in order to prevent their identification. The organoleptic characteristics and presentations of vaccine and placebo were identical. Therefore, the decision to accept or reject a participant was made, and informed consent was obtained from the participant, in ignorance of the assignment in the sequence. The trial participants, investigators, and monitors, were unaware of the trial-group assignments during the whole trial. Statistical analyses were done without knowledge of the groups’ identity. This was known after the analyses were concluded.

### Procedures

The products (Abdala vaccine of placebo) were applied intramuscularly, 0·5 mL in the deltoid region, in a three dose immunization schedule at 0-14-28 days. Concomitant treatment was not anticipated.

Confirmed cases of COVID-19 were defined in participants with a nasopharyngeal swab positive for SARS-CoV-2 by reverse-transcriptase polymerase chain reaction (RT-PCR), who presented at least one major symptom or sign (new onset dyspnoea or worsening, oxygen saturation [SpO_2_] ≤92% by pulse oximetry without oxygen supplement, persistence of chest pain, change of behaviour or alteration in the state of consciousness, local or generalized cyanosis or pneumonia by clinical or imaging diagnosis) or at least two minor symptoms or signs (fever ≥38°C, headache, chills, odynophagia, myalgia, fatigue that interferes with daily activity, vomiting and/or diarrhoea, anosmia and/or ageusia).

Details of the trial procedures are provided in the trial protocol. Once the subject was reported as SARS-CoV-2 positive by the Provincial Centres of Hygiene, Epidemiology and Microbiology involved in the study, the result was sent to the clinical sites participating in the Abdala trial where the subjects belonged, resulting in an alert for the research team. Besides that, all subjects had an identity card, with an emergency 24 hours telephone number to contact the main investigator. All the cases were immediately sent to the “Joaquín Castillo Duany” Hospital in Santiago de Cuba, as the centralized care unit for all the COVID-19 patients in the trial, who remained hospitalized for better follow-up, in all the cases. Clinical data were judged by an independent adjudication committee of medical doctors at the hospital, that was unaware to which study group was the patient assigned. Clinical forms of symptomatic disease due to COVID-19 were classified into four categories: mild, moderate, serious and critical disease, following the same internationally criteria used when addressing the clinical spectrum of SARS-CoV-2 infection.^6^

Blood samples were taken to all the included participants at basal time and were evaluated using the validated UMELISA ANTI SARS-CoV-2 (Immunoassay Centre, Havana) which is a qualitative assay for the detection of total antibodies against SARS-CoV-2 in human serum or plasma. Lab analysts’ carried out the test in a blinded fashion. This evaluation was used in order to exclude all the participants with a positive test for the efficacy endpoints analyses.

The adverse events (AE) were carefully registered according the type, duration, severity, outcome and causality relationship. The severity of the adverse events was classified in three levels: (a) mild, if no therapy was necessary; (b) moderate, if a specific treatment was needed, and (c) severe, when hospitalisation or its prolongation was required, the reaction was life-threatening or contributed to patient’s death. A qualitative assessment was used to classify the causal relationship as definite, probable, possible or doubtful.^7^ Adverse reactions (AR) expected with vaccination were especially sought (pain at the injection site, erythema, induration, headache, fever, among others).

All participants were evaluated in the first hour after the administration of each dose by anamnesis, vital signs (temperature, blood pressure, respiratory and cardiac frequencies), inspection of the inoculation site (for the detection of local symptoms) as well as general physical examination. Subsequently, an active / passive surveillance of AE was in place for the 14 days period between each dose, combining home visits of family physicians at the community level (within 72 hours after each dose) with the self-report of the participating of any AE that could occur during all the trial.

### Outcomes

The primary endpoint of the trial was to evaluate the efficacy of Abdala vaccine in preventing a first occurrence of symptomatic COVID-19, in individuals without evidence of prior exposure to SAR-CoV-2 infection with onset 14 days after the third dose of the immunization schedule (in “per protocol” [PP] population). The consistency of vaccine efficacy was evaluated across various subgroups, including sex, age groups, ethnicity and comorbidities Secondary endpoints were the prevention of mild, moderate, serious and critical forms of COVID-19 (in PP population) as well as the prevention of symptomatic COVID-19 cases that occurred 14 days after two doses (in the “modified intention to treat” [mITT] population). Safety outcome included the collection of adverse events in all enrolled volunteers that received at least one dose of the vaccine or placebo (“intention to treat” [ITT] population).

### Statistical analysis

Sample size was calculated considering the total number of cases needed to demonstrate Abdala’s vaccine efficacy (VE) to prevent symptomatic COVID-19. Assuming proportional risks over time with 1:1 randomization of vaccine and placebo, a total of 151 cases of COVID-19 will be needed to detect a 60% reduction in the hazard rate (i.e., 60% of vaccine efficacy) with the lower limit of 95% confidence interval (CI) of 30% to reject the null hypothesis H_0_: VE ≤ 30% with a statistical power of 0·90. Based on the estimated incidence rate of 0·286% in 2 months for the placebo arm in the study sites, the enrolment of 48000 participants were required, anticipating a 2% dropout rate.

Two interim analyses were planned when approximately 35% and 70% of the target total number of cases (151) were observed with a one-sided O’Brien-Fleming boundary for efficacy. In the interim analysis, the Lan-DeMets spending function was used to control the type I error to be within a 1-sided α=0·025. There was no intention to stop the trial if early efficacy was demonstrated during interim analyses. Details of the sample size calculation are shown in the protocol (Appendix).

In each analysis, survival functions of the control and experimental groups were estimated using the Kaplan-Meier method that allowed a non-parametric estimation of the survival function from censored data. Participants had their data censored at the end of their follow-up. Graphs of the cumulative risk functions were obtained. Log-rank hypothesis tests were performed to check for differences between the survivals curves of the study groups. Vaccine efficacy was defined as the percentage reduction in the hazard ratio for the primary end point (Abdala vaccine vs. placebo). VE was calculated by the expression VE=1-HR, where HR represents the ratio between the risk functions of the experimental and control groups. This ratio was estimated by Cox regression models for proportional hazards. The validation of the Cox model was done by analysing their respective residual graphs and testing their Schoenfeld residuals.

To assess safety, AE were tabulated and plotted by study group. Incidence rates of AE in each group were described in the ITT population defined as the total cohort of participants who received at least one injection, and considered in the group where they were randomized.

The mITT population was defined as the subset of individuals in the ITT definition that showed no previous immunologic or virological evidence of COVID-19 infection at the time of study inclusion, prior to the first dose of investigational product. This population was used for the secondary outcome of efficacy after only two vaccine doses.

The primary efficacy end point in the interim and primary analyses was assessed in the PP population defined as the subset of volunteers included, who met all the eligibility criteria, with no previous immunologic or virological evidence of COVID-19 infection at the time of study inclusion, prior to the first dose of investigational product, who received the complete three dose immunization schedule, in whom the primary endpoint assessment was available and who did not have any major protocol deviations. Individuals were considered in the group where they were randomized. Subgroup analyses were performed by baseline characteristics, including age, sex, ethnicity and presence or absence of coexisting conditions, also carried out in the PP population. Statistical analyses were conducted by independent statisticians using R version 3.6.2.

This study was registered with Cuban Public Clinical Trial Registry, RPCEC00000359.

### Role of the funding source

The funder of the study had a role in study design, data interpretation, and writing of the report, but had no role in data collection or data analysis.

## Results

From 22 March 2021 to 03 April 2021 a total of 48290 subjects were included out of 49656 that were screened. Their disposition is shown in Figure 1. The subjects were randomly distributed into two study groups of the vaccination schedule (0-14- 28 days): placebo and Abdala vaccine (RBD 50 μg). All volunteers completed the vaccination schedule (three doses), except for 1131 subjects that discontinued vaccination (542 in the placebo group, and 589 in the group receiving the Abdala vaccine). Figure 1 also summarizes the number of individuals, by cause, who discontinued the 2nd and 3rd application of the investigational product. The main cause of withdrawal was voluntary dropout which occurred in 61·5% (695/1131) of the subjects. Other causes of discontinuation were: SARS-CoV-2 infection, non-adherence to the protocol by subjects (optional decision provided in the research protocol), decompensation of chronic diseases, among others. During the vaccination period 10 subjects died (five in each study group) from causes not attributable to the investigational product (see Figure 1). Considering all subjects included, high compliance with the planned vaccination was observed (97·7%).

**Figure 1.**
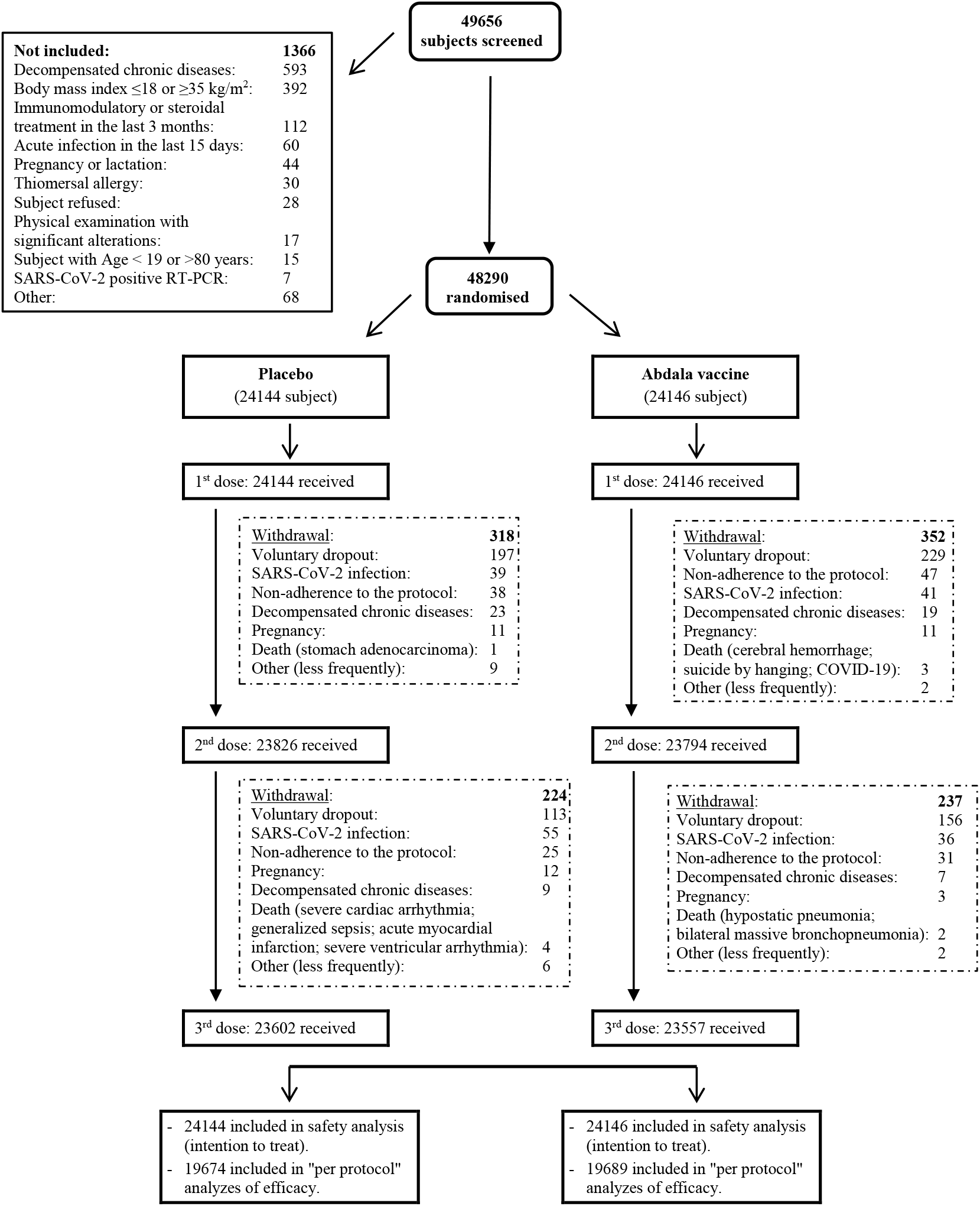
Trial profile.

Table 1 shows the demographic and baseline characteristics of the subjects. The majority of participants were females (52·4%) and the mean (±SD) age was 48·9 ± 16·2 years. Overall, 52·2% of the participants were mestizo, 26·1% were white and 21·7% were black, according to the ethnic distribution of the Cuban population in the south eastern region of the country. No relevant imbalances between the study groups were observed.

**Table 1.** Demographic and baseline characteristics of the participants in the Abdala-3 trial at enrollment.

| Variable |  | Placebo | Abdala vaccine | Total |
| --- | --- | --- | --- | --- |
| N (%) |  | 24144 (50.0) | 24146 (50.0) | 48290 (100) |
| Sex – no. (%) | Female | 12689 (52.6) | 12597 (52.2) | 25286 (52.4) |
|  | Male | 11455 (47.4) | 11549 (47.8) | 23004 (47.6) |
| Age | years | 48.9 ± 16.2 | 48.9 ± 16.1 | 48.9 ± 16.2 |
| Ethnicity<br>– no. (%) | White | 6248 (25.9) | 6357 (26.3) | 12605 (26.1) |
|  | Black | 5233 (21.7) | 5266 (21.8) | 10499 (21.7) |
|  | Mestizo | 12633 (52.3) | 12523 (51.9) | 25186 (52.2) |
| BMI | Kg/m<sup>2</sup> | 26.5 ± 4.2 | 26.5 ± 4.2 | 26.5 ± 4.2 |
| Subjects with some comorbidity – no. (%) |  | 17928 (74.3) | 17931 (74.3) | 35859 (74.3) |
| Main comorbidities / risk factors – no. (%) |  |  |  |  |
| Overweight / type I obesity |  | 11806 (48.9) | 11756 (48.7) | 23562 (48.8) |
| High blood pressure |  | 8223 (34.1) | 8261 (34.2) | 16484 (34.1) |
| Smoking |  | 6339 (26.3) | 6197 (25.7) | 12536 (26.0) |
| Diabetes mellitus |  | 1959 (8.1) | 1945 (8.1) | 3904 (8.1) |
| Bronchial asthma |  | 1881 (7.8) | 1928 (8.0) | 3809 (7.9) |
| Heart disease |  | 897 (3.7) | 897 (3.7) | 1794 (3.7) |
| Other CNS and peripheral disorders |  | 878 (3.6) | 899 (3.7) | 1777 (3.7) |
| Allergy |  | 852 (3.5) | 853 (3.5) | 1705 (3.5) |
| Gastrointestinal disorders |  | 789 (3.3) | 799 (3.3) | 1588 (3.3) |
| COPD |  | 259 (1.1) | 243 (1.0) | 502 (1.0) |
| Cancer |  | 250 (1.0) | 227 (0.9) | 477 (1.0) |
| Ischemic / hemorrhagic cerebral infarction |  | 151 (0.6) | 163 (0.7) | 314 (0.7) |
| Genitourinary disorders |  | 149 (0.6) | 132 (0.5) | 281 (0.6) |
| Disorders of red blood cells and platelets |  | 85 (0.4) | 82 (0.3) | 167 (0.3) |
| Deep and superficial vein thrombosis |  | 28 (0.1) | 36 (0.1) | 64 (0.1) |

Plus-minus values are means ± SD.
BMI: Body-mass index (is the weight in kilograms divided by the square of the height in meters. The calculation was based on the weight and height measured at the time of screening). RBD: receptor binding domain.
CNS: Central Nervous System; COPD: Chronic Obstructive Pulmonary Disease

Seventy-four percent of subjects had some comorbidity or risk factor, with a similar distribution in both study groups. The most frequent were overweight / type I obesity, hypertension, smoking, diabetes mellitus and bronchial asthma, being of interest due to their clinical relevance in relation to COVID-19 (Table 1). The population involved in this Abdala vaccine efficacy clinical trial was very closed to real-world conditions.

The Abdala vaccine was well tolerated. The overall incidence of AR was 446/24144 (1·9%) and 615/24146 (2·5%) individuals in the placebo and Abdala vaccine group, respectively. Reactogenicity was largely absent or mild in most reports, and the following doses were neither withheld nor delayed due to reactogenicity (Figure 2).

**Figure 2.**
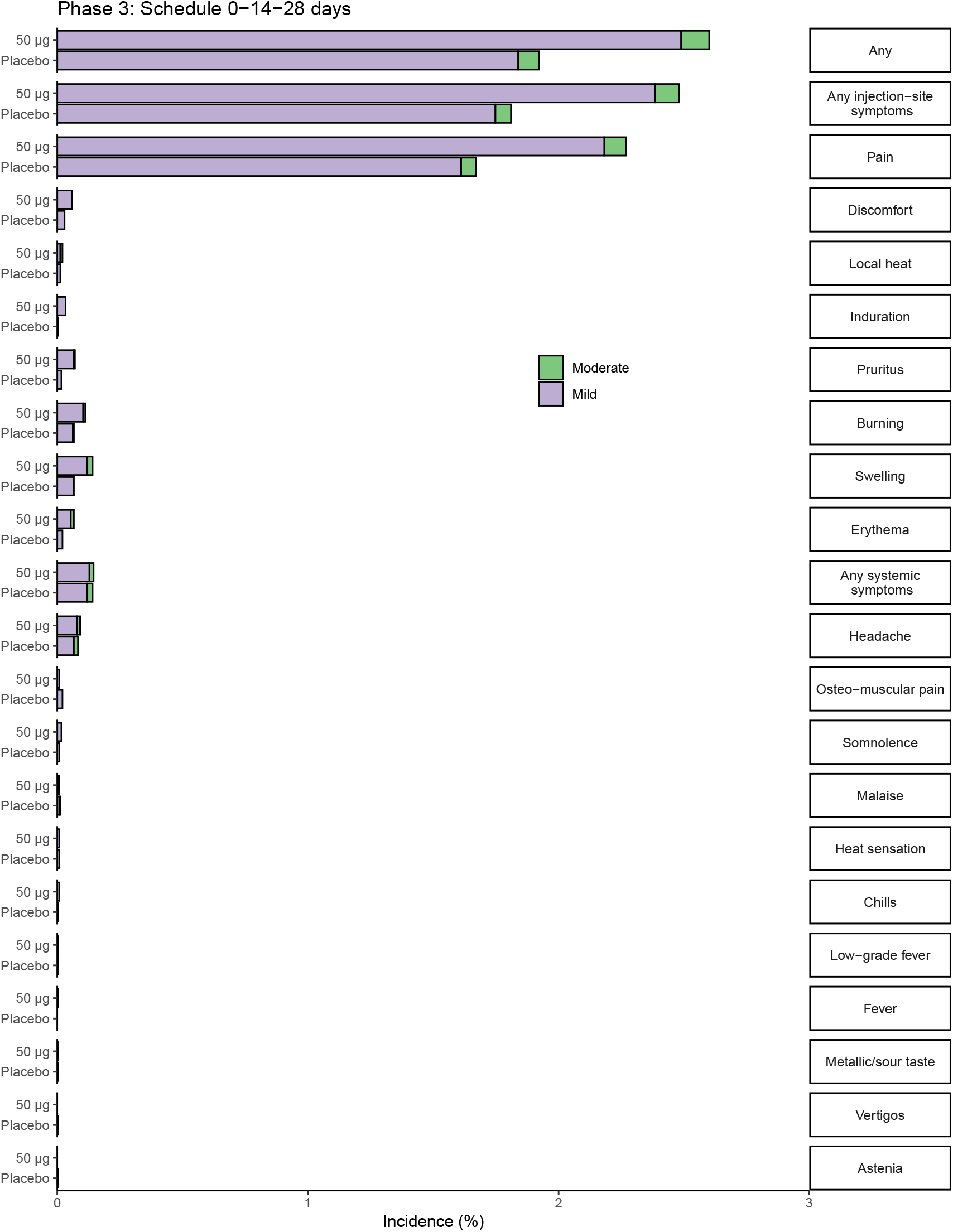
Percentage of participants according to the occurrence of adverse reactions, by study group. The percentage of participants in each study group (Abdala vaccine, Placebo) with adverse reactions according to the maximum FDA (Food and Drug Administration) toxicity grade (mild or moderate) from first dose up to 14 days after third dose is plotted by signs or symptoms. Participants who reported 0 events make up the remainder of the 100%.

Depending on the number of doses applied, reactogenicity was very low: 509 events in 71572 doses applied (0·7%) in the placebo group, and 733/71497 (1·0%) in the group that received Abdala vaccination, with a huge predominance (>90%) of local AR. These reports were similar between study groups, slightly higher in the group receiving the Abdala vaccine. More than half of these AR were reported during the observation period corresponding to the first dose, being significantly reduced with the following applications. Most of the AR resolved spontaneously in the first 24-48 hours without medication.

No severe AE with demonstrated cause-effect relationship attributable to the investigational product were reported and there were no withdrawals for this cause. No episodes of anaphylaxis, vaccine-associated enhanced COVID-19 or other immune-mediated medical conditions, myocarditis as well as other events of special interest relevant to COVID-19 were reported in this investigation.

During the vaccination period, a total of 10 serious AE leading to death occurred in five participants in the placebo group, and five subjects in the Abdala vaccine group, described in Figure 1. No deaths were considered by investigators to be related to the vaccine or placebo. Subsequently, during the evaluation of the vaccine efficacy (from 14 days after completion of the vaccination schedule), there were four deaths related to COVID-19, all in the placebo group.

The efficacy results against symptomatic COVID-19 in the first interim analysis with 53 COVID-19 cases (50 in the placebo group and three in the vaccine group) was 94·00% (95% CI 80·75-98·13). In the second analysis carried-out when 108 cases were accumulated (101 in placebo group and 7 in the vaccine group) the efficacy was of 93·08% (85·11-96·78%).

Finally, in the per-protocol population of 39363 participants, 153 cases of virologically confirmed symptomatic COVID-19 with an onset at least 14 days after the third dose occurred: 142 in placebo recipients and 11 in Abdala vaccine recipients. This corresponded with a vaccine efficacy of 92·28% (95% CI 85·74-95·82) that was the primary end-point of the trial (Figure 3).

**Figure 3.**
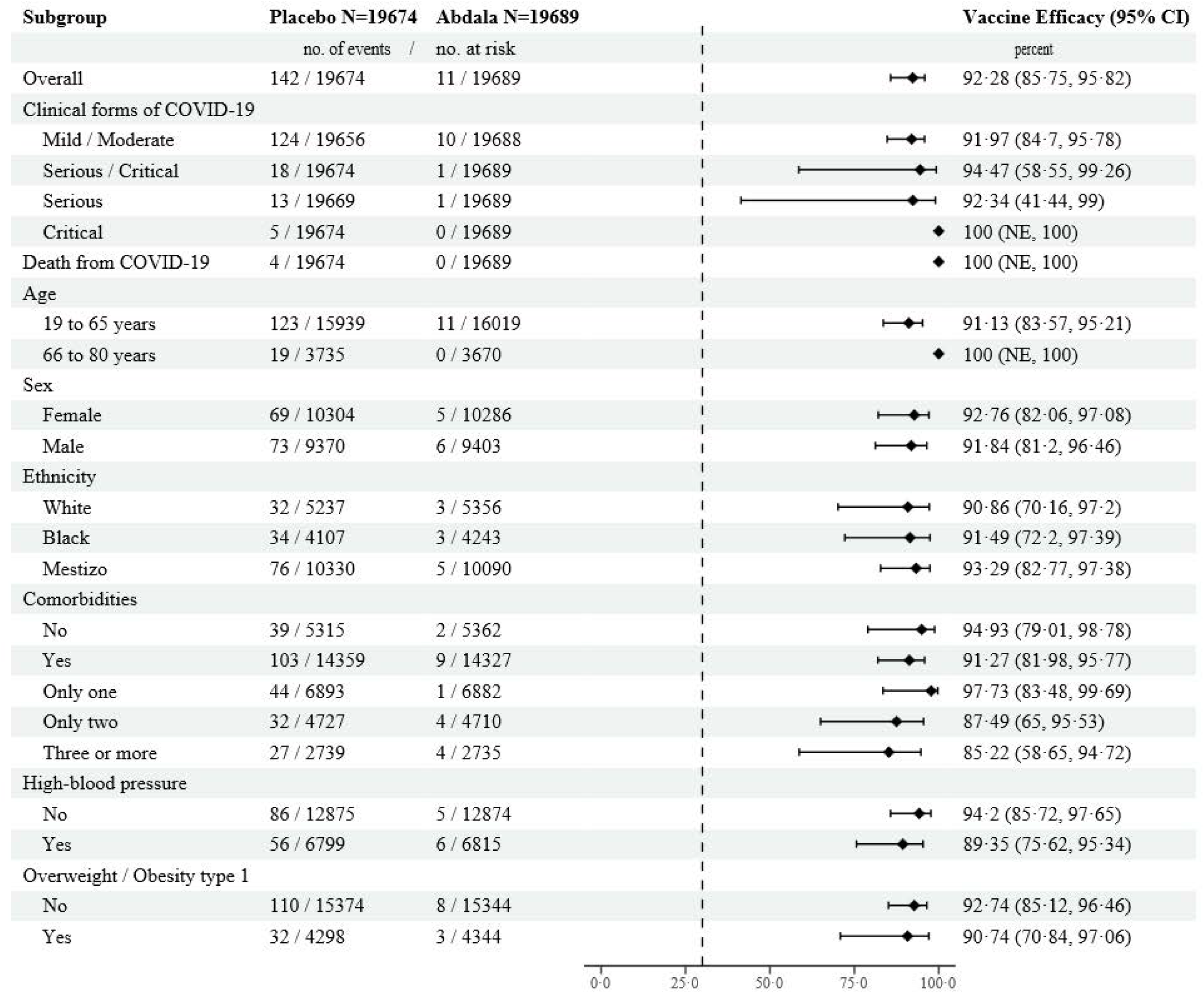
Vaccine Efficacy of Abdala vaccine to Prevent Symptomatic COVID-19. Secondary end-points and Subgroups analyses. Analysis of the efficacy of Abdala vaccine in the prevention of symptomatic COVID-19 by clinical forms of the disease, death and various subgroups in the per-protocol population, was based on adjudicated assessments starting 14 days after the third injection. Vaccine efficacy, defined as 1 minus the hazard ratio (Abdala vs. placebo), and 95% confidence intervals were estimated with the use of a stratified Cox proportional-hazards model. The dashed vertical line represents a vaccine efficacy of 30%, based on the null hypothesis that the primary efficacy of Abdala vaccine is 30% or less

The cumulative incidences of COVID-19 related events in the vaccine and placebo groups are shown in Figure 4.

**Figure 4.**
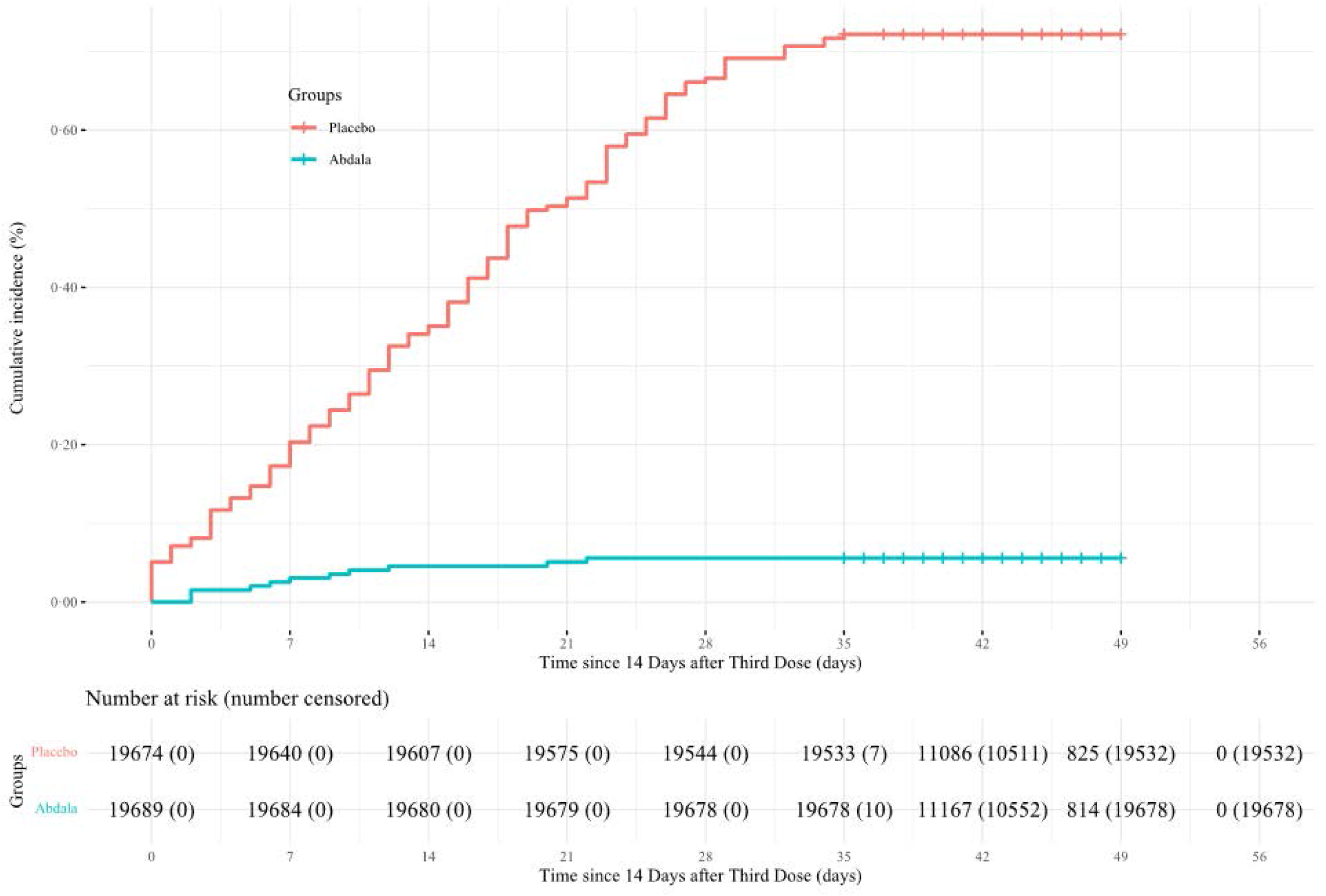
Kaplan-Meier Plots of Efficacy of Abdala vaccine against Symptomatic COVID-19. Shown is the cumulative incidence of symptomatic Covid-19 in the per-protocol population. The timing of surveillance for symptomatic COVID-19 began at least 14 days after the administration of the third dose (i.e., on day 42) through approximately the first 2 months of follow-up.

A secondary endpoint was the evaluation of the efficacy according to the clinical forms of COVID-19. In the case of mild/moderate forms (124 cases in the placebo group and 10 in vaccine group) the vaccine efficacy was 91·96% (95% CI 84·69-95·78). The most severe forms of COVID-19 (serious/critical disease) occurred in 19 participants (18 in the placebo group and only one in the vaccine group) for a vaccine efficacy of 94·46% (58·52-99·28). The five patients critically ill occurred only in placebo recipients, as well as the four deaths, indicating that Abdala vaccine efficacy for both conditions was of 100% (95% CI could not be estimated to 1·0) (Figure 3).

A secondary analysis of symptomatic cases after the administration of only the first two doses of the schedule was carried out in the mITT population (from which individuals who has discontinue the trial, for any reason, before the second dose or who had an asymptomatic infection in that period were eliminated). Among the 19885 participants in the placebo group 41 COVID-19 events were found *vs*.7 out of 19890 participants in the vaccine group. This corresponded with a vaccine efficacy of 82·96% (95% CI 62·01-92·35) for the prevention of COVID-19 after the administration of only two doses of vaccine.

Additional analyses of vaccine efficacy in preventing COVID-19 were performed in subgroups defined by age, sex, and ethnicity, as well as the presence or absence of coexisting conditions (Figure 3). In the age group of 66 to 80 years, no cases of COVID-19 were found in those who received the Abdala vs. 19 in the placebo group.

Vaccine efficacy by sex and ethnicity was above 90%. In participants with/without comorbidities the efficacy was of 91·27% (95% CI 81·98-95·77) and 94·93% (79·01-98·72), respectively. The point estimated of efficacy decreased as the number of comorbidities increased. Finally the two most frequent comorbidities founded in the trial (hypertension and overweight / obesity type 1) were analysed separately showing efficacy estimates also above 90% (Figure 3). The majority of symptomatic cases in the efficacy analyses had some comorbidity (103/142 cases in the placebo group and 9/11 for the vaccine group).

## Discussion

Worldwide more than 360 vaccines against SARS-CoV-2 based on a broad range of technological platforms are currently under development, 170 of them already in clinical trials, including 43 in phase 3 and eleven vaccines had been included in the WHO’s emergency authorisation list.^8^ Results of efficacy of different vaccines are already published^9-16^ but the comparison between them has been difficult considering the differences in study protocols, dose regimen, the evaluation in non-equivalent conditions in different countries and economic conditions, various stages of COVID-19 pandemic and SARS-CoV-2 variants, among other factors.^17-19^

A three dose vaccination schedule of Abdala vaccine administered at 0-14-28 days was found to have high efficacy against symptomatic COVID-19 (92·28%), fulfilling the WHO target product profile for COVID-19 vaccines that suggest a “clear demonstration of efficacy (on a population basis) ideally with a point estimate around 50%” should be a minimum acceptable criterion for any COVID-19 vaccine, and as a preferred condition, at least 70% with consistent results in older individuals,^20^ that were both achieved in this phase 3 trial. WHO recommends that successful vaccines should show an estimated hazard reduction of at least half, accurately enough to conclude that the true efficacy of the vaccine is greater than 30%.^20^ U.S. Food and Drug Administration (FDA) guidance also includes this 30% lower limit as criteria for authorization.^21^ It is considered that a vaccine that is 50% effective could significantly reduce the incidence of COVID-19 in vaccinated people and could provide useful collective immunity. Therefore, although a much higher efficiency of 50% would be better, it would represent substantial progress.^22^

The efficacy results against symptomatic COVID-19 in the first and second interim analysis was high, 94·00% and 93·08% respectively, fulfilling the hypothesis that VE >30% in both cases, thus, fulfilling WHO’ requirements for COVID-19 vaccines, were also met since the first analysis.

The efficacy results obtained according to the clinical forms of the symptomatic disease were also encouraging, showing that the vaccine achieved an efficacy of 91·96% for mild/moderate disease and most important, 94·46% for serious/critically ill patients. No patient in the vaccine group was classified as critical and no deaths occurred in that group (100% efficacy, with no estimated lower confidence intervals).

Although this trial was not powered to definitively assess efficacy by subgroups, the point estimates of efficacy according age, sex, ethnicity, as well as the presence of comorbidities were also high, above 90%, consistent with the primary efficacy end-point. In all the cases the lower limit of the 95% confidence interval was higher than 30%. Considering that mortality from SARS-CoV-2 disproportionately affects older adults, it is very important to develop vaccination strategies to protect this highly vulnerable subgroup. For that reason we considered the enrolment in the trial of participants from 66-80 years of age. Interestingly, in the PP evaluation of the efficacy in this group of age, none case of symptomatic disease was found in the vaccine group (0/3770 participants), but 19 cases out of 3775 subjects were reported in the placebo group.

Taken into account the elevated proportion of participants that suffered from pre-existing conditions, one the most relevant subgroup analyses was that of comorbidities. When the absence or presence of any comorbidity was analysed, the efficacy values were again around of higher than 90%. It is interesting that the majority of symptomatic cases in the efficacy analyses had at least one comorbidity (72·5% of the cases in the placebo group and even higher, 81·8%, in the vaccine group). The point estimates of efficacy are lower when the number of comorbidities increased. Even in the group of individuals with 3 or more comorbidities, the values obtained exceed the minimum efficacy required by the WHO. Hypertension and overweight/type I obesity were analysed as independent comorbidities, considering that they were the most frequently found, in accordance with the occurrence in real word settings in Cuba. The efficacy results were consistent with the other analysis.

All of the efficacy results presented are relevant, due to the influence of these control variables on the clinical outcome of COVID- 19 patients, confirmed by different groups that found that severe forms of COVID-19 in adults have been associated with the presence of pre-existing medical conditions. Hazard associated with a condition and hospital mortality increased markedly with age.^23-25^ The absolute risk of death from COVID-19 increased by 14%, 11%, and 12% for diabetes, hypertension and obesity, respectively, contributing to nearly 30% of COVID-19 deaths.^26^ BMI may play an important role in COVID-19 and there is a linear dose-response association between BMI, severity, and COVID-19 mortality. In addition, obesity (BMI ≥30 kg/m^2^) was associated with a significantly higher risk of critical COVID-19 and in-hospital mortality.^27^

The efficacy after only two doses was rather high (82·96%), that could be a relevant data for the immunization programs due that individuals may not complete the immunization schedule. However, the three dose schedule is maintained as the recommended one, considering that can be completed in only one month period, achieving a higher efficacy and an expected of longer duration.

To our knowledge, Abdala’s study is one of the largest phase 3 trials published on COVID-19, enrolling more than 48000 people in less than 14 days. The trial performed well, without deviations and a minimum of dropouts (1·4%). This was also facilitated by the compact immunization schedule of three doses applied at 0-14-28 days, previously selected in the phase 2 trial, very useful to face the urgency of the pandemic situation.^5^ Randomisation assured a homogenous demographic and baseline characteristics of the participants in both placebo and vaccine groups. Blinding was maintained along all the trial up to the estimation of clinical efficacy. Therefore, the internal validity of the study was adequate. Several factors such as the inclusion of participants from both sexes and different ethnicity groups, the wide range of ages from 19 to 80 years, the fact that 74% of the study population reported previous comorbidities/risk factors as well as the high sample size of the trial contributes to the generalisability (external validity, applicability) of the trial findings, narrowing the gap between the clinical trial and real world populations, in which the same immunization schedule was applied.

The efficacy evaluation of Abdala vaccine was designed in individuals with no previous exposure to SARS-CoV-2, considering that it was not known if previous infection could provide some kind of protection against subsequent one. For that reason, although the information about known previous COVID-19 was considered as an exclusion criteria, the evaluation of a serum sample of all included individual was performed at time cero (before the administration of the first dose), in order to exclude those positive individuals from the efficacy end-point, but that still would contributed to the safety’s evaluation of the vaccine.

The excellent safety profile observed in the phase 1-2 clinical trial was confirmed in the present phase 3 study.^5^ The Abdala vaccine, in its short three-dose regimen, was well tolerated, without any serious toxicity, and no deaths attributed to his administration. The adverse reactions reported were minimal, mostly mild and from the injection site, of short duration, resolved spontaneously. Injection-site pain was the most common local symptom, followed by burning, swelling, and itching. The local and systemic adverse events after Abdala vaccine administration were comparable with the placebo group, indicating that there was no increased risk of adverse events with the vaccine.

The long-term follow-up of study participants was foreseen for one year, to assess the duration of protection and the identification of potential adverse events such as vaccine-associated enhanced respiratory disease that was not found during the trial. These further follow-up also included those volunteers that received placebo (in the trial) and were further vaccinated with Abdala (after the study finished and known the results) as part vaccination policies established in the country from Cuban’s Ministry of Health, in the course of COVID-19 emergency. To complete this objective data management of safety and immunological information are still on-going and will be part of a further presentation.

In Cuba three epidemic periods have been defined associated with the circulation of three major variants. Our phase 3 clinical trial was conducted from March to June 2021, corresponding exactly with the second period/second wave of the pandemic in the country (between March and June 2021). This period was characterized by a continued increase in the incidence of cases associated with the expansion of the circulation of the Beta variant, and a 11·89-fold increase compared to the total reported cases in 2020 and 3·41 times the total reported from December 2020 to February 2021. The distribution of the variants changed over time according to the emergence and expansion of those with the greatest evolutionary advantage. In March, B.1.1.519 variant was detected and at this time, Beta and D614G circulated with a similar frequency (30·81%). From April to May, an increase in Beta circulation was observed. Additionally, the variants Gamma/P1, B.1.1.523 and Delta (B.1.617.2) were detected for the first time in April. Delta rapidly increased from 1·44% in May to 27·14% in June. In parallel, the Beta variant diminished from 60·58% in May, to 51·26% in June^28^, so, the efficacy of Abdala vaccine was measured in the context of highly circulation of mutant viruses.

Considering the safety and efficacy results achieved, the Cuban regulatory agency (Centre for State Control of Medicines, Equipment and Medical Devices) issued an emergency use authorization (EUA) for Abdala on July 9, 2021 and massive immunization in adult population started.

This study has several limitations. First, the short duration of the efficacy follow-up, as has previously occurred for many other vaccine candidates in Phase 3 trials. It is important to take into consideration that we carried out a unique inclusion strategy, where the 48290 participants were included in less than 14 days, with a uniform immunization schedule of three doses in a 28 days period and a median time of follow-up since randomization up to efficacy data base closure was 75 days (IQR 73, 79 days). On the other hand, as the incidence of SARS-CoV-2 increased in the country with the circulation of VOC, the time required to meet efficacy end-point decreased. Second, although in this report it is not possible to show results on the long-term protective effects of the vaccine during the phase 3 trial, a retrospective cohort study carried out in Havana (Cuba) in 1355638 persons, demonstrated that Abdala vaccine was highly effective in preventing severe illness and death from COVID-19 (primary outcomes of the study) under real-life conditions, during the third epidemic period, where Delta variant prevailed.^29^ Third, this study did not assessed the efficacy in other populations as children, adolescents and pregnant and lactating women, but further studies in those populations were also conducted after EUA was granted. Fourth, in the current report data regarding neutralizing antibodies titters and other immunological variables are not included. However, as part of the evaluation of duration of the response randomly selected serum samples are under analysis, including neutralization against VOC Omicron.

To our knowledge, Abdala is the first COVID-19 vaccine based on a subunit RBD-protein obtained by recombinant-DNA technology in the yeast *Pichia pastoris*, which demonstrated clinical efficacy in a phase 3 clinical trial. Abdala vaccine is safe and highly protective against symptomatic COVID-19, including the most severe forms of the disease and death. Those results, along with its immunization schedule and the advantage of easy storage and handling conditions at 2-8°C, make this vaccine an option for the use in massive immunization strategies as a key tool for the control of the pandemic.

## Supporting information

Appendix - Other investigators

## Data Availability

All data produced in the present study are available upon reasonable request to the authors.

## Contributors

FHB concept and designed this study, was its main coordinator, and took part in the analyses and interpretation of the results and paper writing; MCRC, ZNR, MPM, JBA, MCMR, LLL, VMVP, JSD, YBG, SLV, TMH, NCC and ERM were clinical investigators of the trial, participated in volunteers recruitment, data acquisition and interpretation of the results; YMB, KlUP, KaUP, JQG, IBC, ABI, APM, RMB, DSC and KMCA took part in trial coordination and data monitoring; ERM, COCC, MAV and RHR participated in the data management, statistical analyses and results interpretation; JLRR and GMS ensured masking of the investigational product and the cold chain throughout the process, as well as compliance with GCPs during vaccination; MVCN contributed to the diagnosis and virological follow-up by PCR-RT of patients with COVID-19; MAA and MLF took part in the trial design and coordination; VLMG participated in the trial design, advice, analyses and interpretation of the results and paper writing. All authors had full access to and verify all the data in the study, and took the decision to submit the paper for publication. The authors FHB, MCRC, YMB, ERM, KlUP, KaUP, JQG, COCC, MAV, KMCA and VLMG, verified the study data.

## Declaration of interests

Authors FHB, YMB, KlUP, KaUP, JQG, JLRR, IBC, GMS, MLF, MAV, RHR, KMCA, MAA, and VLMG, are employees of the Centre for Genetic Engineering and Biotechnology, Havana Network, where Abdala vaccine active ingredient is produced and the formulation was developed. The remaining authors have no conflict of interests. No honoraria, consulting fees or payments for seminar presentations, speeches or appearances have been received by any of the authors.

## Data sharing

The study protocol is provided in the appendix. Anonymised participant data will be made available when the trial is complete, upon requests directed to the corresponding author. Proposals will be reviewed and approved by the sponsor, investigator, and collaborators on the basis of scientific merit. After approval of a proposal, data can be shared through a secure online platform after signing a data access agreement.

## Acknowledgements

A special thanks to the thousands of volunteers in this trial, who maintained the highest adherence to the research protocol. Likewise, the authors wish to thank the government authorities of the provinces involved in the study, and the Cuban Ministry of Public Health for their support.

## Notes

### Clinical Trial

RPCEC00000359

### Author Declarations

ad hoc Centralized Ethics Committee, integrated by members of the Ethics and Review Committee of the Provincial Hospital Saturnino Lora in Santiago de Cuba (main clinical site), extended with members of the Research Ethics Committees of the Universities of Medical Sciences of Santiago de Cuba, Guantanamo and Granma, gave ethical approval for this work.

## References

1. World Health Organization. Weekly epidemiological update on COVID-19 - 24 August 2022. Available at: https://www.who.int/publications/m/item/weekly-epidemiological-update-on-covid-19 24-august-2022. Accessed August 27, 2022.

2. Centers for Disease Control and Prevention. COVID-19 Risks and Vaccine Information for Older Adults. Available at: https://espanol.cdc.gov/coronavirus/2019-ncov/need-extra-precautions/older-adults.html. accessed March 14, 2022.

3. Limonta-Fernández M, Chinea-Santiago G, Martín-Dunn AM, et al. An engineered SARS-CoV-2 receptor-binding domain produced in Pichia pastoris as a candidate vaccine antigen, New BIOTECHNOLOGY 2021. doi: 10.1016/j.nbt.2022.08.002

4. Izquierdo M, Ramos Y, Costa L, et al. Demonstrating “Abdala” subunit vaccine thermostability. BioProcess J 2022;21. doi: 10.10.12665/J21OA.Izquierdo

5. Hernández-Bernal F, Ricardo-Cobas MC, Martin-Bauta Y, et al. Safety, tolerability, and immunogenicity of a SARS-CoV-2 recombinant spike RBD protein vaccine: a randomised, double-blind, placebo-controlled, phase 1-2 clinical trial (ABDALA Study). eClinicalMedicine 2022;46:101383. doi: 10.1016/j.eclinm.2022.101383

6. National Institutes of Health (NIH). Coronavirus Disease 2019 (COVID-19). Treatment Guidelines Panel 2021. Available in: https://www.covid19treatmentguidelines.nih.gov/. accessed March 10, 2021.

7. Naranjo CA, Shear NH, Busto U. Adverse drug reactions. In Kalant H and Roschlau WHE, eds. Principles of medical pharmacology. Oxford University Press, New York 1998:791-800.

8. COVID-19 vaccine tracker and landscape. https://www.who.int/publications/m/item/draft-landscape-of-covid-19-candidate-vaccines (accessed August 31, 2022).

9. Baden LR, El Sahly HM, Essink B, et al. Efficacy and safety of the mRNA-1273 SARS-CoV-2 vaccine. N Engl J Med 2021; 384(5):403–16. doi: 10.1056/NEJMoa2035389.

10. Polack FP, Thomas SJ, Kitchin N, et al. Safety and efficacy of the BNT162b2 mRNA COVID-19 vaccine. N Engl J Med 2020;383:2603–15. doi: 10.1056/NEJMoa2034577.

11. Voysey M, Costa SA, Madhi SA. et al. Single-dose administration and the influence of the timing of the booster dose on immunogenicity and efficacy of ChAdOx1 nCoV-19 (AZD1222) vaccine: a pooled analysis of four randomised trials. Lancet 2021;397:881–91. https://doi.org/10.1016/S0140-6736(21)00432-3.

12. Sadoff J, Gray G, Vandebosch A, et al. Safety and efficacy of single dose Ad26.COV2.S vaccine against COVID-19. N Engl J Med 2021;384:2187–201. doi: 10.1056/NEJMoa2101544.

13. Logunov DY, Dolzhikova IV, Shcheblyakov DV, et al. Safety and efficacy of an rAd26 and rAd5 vector-based heterologous prime-boost COVID-19 vaccine: an interim analysis of a randomised controlled phase 3 trial in Russia. Lancet 2021;397:671–81. doi: 10.1016/S0140-6736(21)00234-8.

14. Heath PT, Galiza EP, Baxter DN, et al. Safety and efficacy of NVX-CoV2373 Covid-19 vaccine. N Engl J Med 2021;385(13):1172–83. doi: 10.1056/NEJMoa2107659.

15. Al Kaabi N, Zhang Y, Xia S, et al. Effect of 2 inactivated SARS-CoV-2 vaccines on symptomatic COVID-19 infection in adults: A randomized clinical trial. JAMA 2021;326(1):35–45. doi: 10.1001/jama.2021.8565.

16. Tanriover MD, Doğanay HL, Akova M, et al. Efficacy and safety of an inactivated whole-virion SARS-CoV-2 vaccine (CoronaVac): interim results of a double-blind, randomised, placebo-controlled, phase 3 trial in Turkey. Lancet 2021;398:213-https://doi.org/10.1016/S0140-6736(21)01429-X.

17. Cheng H, Peng Z, Luo W et al. Efficacy and Safety of COVID-19 Vaccines in Phase III Trials: A Meta-Analysis. Vaccines 2021;9(6):582. https://doi.org/10.3390/vaccines9060582.

18. Sharif N, Alzahrani KJ, Ahmed SN, Dey SK. Efficacy, Immunogenicity and Safety of COVID-19 Vaccines: A Systematic Review and Meta-Analysis. Front Immunol 2021;12:714170. doi: 10.3389/fimmu.2021.714170.

19. Rotshild V, Hirsh-Raccah B, Miskin A, Muszkat M, Matok I. Comparing the clinical efficacy of COVID-19 vaccines: a systematic review and network meta_analysis. Sci Rep 2021;11:22777. https://doi.org/10.1038/s41598-021-02321-z.

20. World Health Organization. Target product profiles for COVID-19 vaccines. April 9, 2020. Available at: https://www.who.int/who-documents-detail/who-target-product-profiles-for-covid-19-vaccines. accessed July 17, 2022.

21. US Food and Drug Administration. Development and licensure of vaccines to prevent COVID-19: guidance for industry. June, 2020. Available at: https://www.fda.gov/regulatory-information/search-fda-guidance-documents/development-and-licensure-vaccines-prevent-covid-19. accessed December 12, 2021.

22. Krause P, Fleming TR, Longini I, Henao-Restrepo AM, Peto R, World Health Organization Solidarity Vaccines Trial Expert Group. COVID-19 vaccine trials should seek worthwhile efficacy. Lancet 2020; 396(10253):741–3. doi: 10.1016/S0140-6736(20)31821-3.

23. Kompaniyets L, Pennington AF, Goodman AB, et al. Underlying Medical Conditions and Severe Illness Among 540,667 Adults Hospitalized With COVID-19, March 2020–March 2021. Prev Chronic Dis 2021;18:210123. http://dx.doi.org/10.5888/pcd18.210123.

24. Grasselli G, Greco M, Zanella A, et al. Risk Factors Associated With Mortality Among Patients With COVID-19 in Intensive Care Units in Lombardy, Italy. JAMA Intern Med 2020;180(10):1345–55. doi: 10.1001/jamainternmed.2020.3539. Erratum in: JAMA Intern Med 2021;181(7):1021. doi: 10.1001/jamainternmed.2021.1229.

25. Kim L, Garg S, O’Halloran A, et al. Risk Factors for Intensive Care Unit Admission and In-hospital Mortality Among Hospitalized Adults Identified through the US Coronavirus Disease 2019 (COVID-19)-Associated Hospitalization Surveillance Network (COVID-NET). Clin Infect Dis 2021;72(9):e206–14. doi: 10.1093/cid/ciaa1012.

26. Mahamat-Saleh Y, Fiolet T, Rebeaud ME, et al. Diabetes, hypertension, body mass index, smoking and COVID-19-related mortality: a systematic review and meta-analysis of observational studies. BMJ Open 2021;11(10):e052777. doi: 10.1136/bmjopen-2021-052777. PMID: 34697120; PMCID: PMC8557249.

27. Du Y, Lv Y, Zha W, Zhou N, Hong X. Association of body mass index (BMI) with critical COVID-19 and in-hospital mortality: A dose-response meta-analysis. Metabolism 2021;117:154373. doi: 10.1016/j.metabol.2020.154373.

28. Guzmán MG, Pérez L, Tejero Y, et al. Emergence and evolution of SARS-CoV-2 genetic variants during the Cuban epidemic. J Clin Virol Plus 2022; 2(4):100104. doi: 10.1016/j.jcvp.2022.100104.

29. Más-Bermejo PI, Dickinson-Meneses FO, Almenares-Rodríguez K, et al Cuban Abdala Vaccine: Effectiveness in Preventing Severe Disease and Death from COVID-19 in Havana, Cuba; a Cohort Study. The Lancet Regional Health - Americas 2022. doi: 10.2139/ssrn.4072478.

