## Appendix - Other investigators for "A phase 3, randomised, double-blind, placebo-controlled clinical trial for adult evaluation of the efficacy and safety of a SARS-CoV-2 recombinant spike RBD protein vaccine (ABDALA-3 Study)"

**“Saturnino Lora” Hospital, Santiago de Cuba** (E Colina-Ávila MD, J Rodríguez-Nuviola MD, Y Naranjo-Vargas MSc, A Rosado-Rosado MSc, S del Valle-Piñera MD, L del Toro-Lahera MD, L Mantejo-Hormigó MD, M Ramírez-Domínguez MD, M A Vinent-Terazón MD, J Manuel-Fernández MD, D Ramis-Rosales MD, D Ávila-Fuentes MD, E Tablada-Ferreiro MD, L Aguilera-Ramos MD, M C Acosta-Portuondo MD, N A Ali-Mros MD, Y Álvarez-Calvo BSc, V Ruano-González MSc, R Remedios-Reyes MD, O Poll-Fiss MSc, L Garbey-Delás BSc, J Sigüenza-Castilla BSc, R Suárez-Domínguez MD, L R Castro-Andión MD, M Gay-Muguercia MD, L A Recacén-Canet MSc, S Jaime-Vázquez MSc, M Domínguez-Cardosa MSc, M Clares-Pochet MSc, O Veliz-Aranda MD, E Mayor-Guerra BSc, A Pozo-Estrada MD, T Sanchez-Borot MD, M F Moreno-Soto MD, Y Ferrer-Bonne MSc, E Sánchez-Carbonell BSc, Y Sotomayor-Murgada BSc, I Ramirez-Bicet BSc, E Ramos-Caraballo BSc, P Rodríguez-Reyes BSc, R A Ferrer-Socarrás BSc, S Hernández-Fals BSc, Y Días-Hodelín BSc, H J Pérez-Hernández MD); **“Julián Grimau” Polyclinic, Santiago de Cuba** (I García-Carrión MD, S Semanat-Destrade MD, N Cese-González MD, E M Grajales-Pozo MD, Y Maceo-Masó MD, E Gey-Gayol MD, D Dupon-Sanet MD, Y Babastro-Pérez BSc, R Ramos-González BSc, M Vazquez-Rodríguez BSc, M Viera-Muñiz BSc, S Ferrer-Ramírez BSc); **“Armando García” Polyclinic, Santiago de Cuba (**M L Reyes-López MD, M Herrezuelo-Heredia MD, Z Sarda-Prada MD, H Vidal-Ibarra MD, L M García MD, L A Lopez-Arzuaga MD, M Heredia-Tamayo MD, M Martin-Figueras MD, D Rodríguez-Fernández MD, O R Martínez-Rodríguez MD, A Balanquer-Herrera BSc, D Hernández-Mayeta BSc, I Castillo-Rodríguez BSc, E Rodríguez-Bridón BSc, N Velázquez-Noe BSc, V Leyva-Sorribe BSc); **“José Martí” Polyclinic, Santiago de Cuba** (D González-Menés MD, R Labrada-González MD, Y Beltrán-Matamoros MD, T Machado-Herrera MD, Y Bring-Monteagudo MD, Y Hung- Brido MD, T Marin-Alvarez MD, Y Ramirez-Zamora MD, A Zapata-Torreblanca MD, Y Rivera-Garcia MD, R Lopez-Palay MD, D Chávez-Pérez MD, A Rosales-Canedo MD, A Bizet-Puentes MD, D Blanco-Gómez MD, Y Planas-Linares MD, K Velázquez-Arrue MD, D Fuentes-Rivera MD, Y Robert-García MD, A Hiler-Zamora MD, L Bain-Valiente MD, Y Spek-Valverde MD, E A Fuentes-Despaigne MD, M Savigne-Pérez BSc, I González-Duanys BSc, B Rivera-Elías BSc, I Batista-Lecay BSc, V Parra-Licea BSc, E Llamos-Infante BSc, G Rivera-Elías BSc, H Mas-Ruiz BSc, M Guzmán-Cisnero BSc, Y Macias-Gonzalez BSc, J Villalon-Guillort BSc, T Garlobo-Rodríguez BSc, D Terrero-Benavides BSc, M Casternau-Cedeño BSc, A Martínez-Fajardo BSc, V Y Hanet-López BSc, V Melis-Torres BSc, I M González-Duanys BSc, L Guerra-La O BSc, M Miranda-Beltrán BSc, S Calderín-Figueredo BSc, Y Perez-Jaime BSc, D Mateo-Estei BSc, I Delis-Duany BSc, Y de la Vega-Ramírez BSc, A Moreno-Alcolea BSc, M D La O-Rodríguez BSc, A Betancourt-Betancourt BSc, Y Collazo-Perera BSc, M Ge-Copa BSc); **“Ramón López Peña” Polyclinic, Santiago de Cuba** (M Matos-Fonseca MD, S Hernández-Carrión MD, L B Tellez-Domínguez MD, R Batista-Almaguer MD, A Y Arias-Caballero MD, S Catá-Navarro MD, A Moreno-Venero MD, D O-Connor MD, Y Cobas-Bataille MD, D Venero-Fuentes MD, Y Diaz-Castillo MD, F Cardero-Castillo MD, L R Rodríguez-Delis MD, M Lubín-Sánchez MD, D Isaac-Aranda MD, K F Pacheco-Arias MD, M Riveri-Larduet MD, J A Martínez-Ortiz MD, Y Álvarez-Heredia MD, H A Carbonell-Meriño MD, A E Saiz-Doimeadiós MD, Y Torres-Leyva MD, L Maia-Amaro MD, I López-Ferrer MD, Y Hechavarría-Despaigne MD, A Alonso-Torres MD, A Ramírez-Reyes MD, N L Soriano-Tamayo BSc, Z Dominica-Pérez BSc, T Herrera-Belén BSc, L N Medina-Villalón BSc, L Isaac-Martínez BSc, M Carbonell-Cardona BSc, T Céspedes-Quesada MSc, Y Verdecia-Boris BSc, D C Cupull-Rodríguez MSc , C R Corniel-Suárez BSc, L Isacc-Rodríguez BSc, E Pérez-Martínez BSc, I Duret-Martínez BSc , D A Ferrer-Vargas BSc, L E Muriel-Mustelier BSc, J Bravo-Bravo BSc, I Castro-Hierrezuelo BSc, Y Miguel-Rondon BSc, O Smith-Medina BSc, A R Puron-Oliva BSc, Y Matos-Ramirez BSc, M Miranda-Cabrera BSc, D León-Rodríguez BSc, L A Apure-Rodríguez Tech, Y Carbonell-Nuñez MD, L Vicet-Mandariega MD, R Massant-Prado MD); **“28 de septiembre” Polyclinic, Santiago de Cuba** (Y Larrea-Conte MD**,** N Dussac-Tamayo MD**,** E Díaz-Milian MD**,** Y B Pérez-González MD**,** D Carvajal-Patas MD**,** G Galano-Leyva MD, M del Carmen Mora-Dupuy MD, M Pérez-Rivera MD, A Coca-Sánchez MD, L Gascón-Fernández MD, B D Ferrer-Same MD, E Gey-Mustelier MD, Y Danger-Cuadra MD, Y Fernández-García MD, Y Acosta-Suárez MD, A Higuera-Ellis MD, O García-Céspedes BSc, B Gallardis-Parra BSc, Y Cortes-Olivares BSc, A Mustelier-Tornes BSc, M Bandera-Vinent BSc, M González-Utria BSc, L Pereiro-Salazar BSc, J Paumier-Catanares BSc, C Betancourt-Espinosa BSc, Y Silva-Illas BSc, A Trompeta-Martínez BSc, O Despaigne-Garbey BSc, L Feria-Lugo BSc, Y Hernandez-Delgado MD, A Medina-Rojas MD, A Issac-Balón Tech, A M Leyva-Sotomayor Tech); **“Frank País” Polyclinic, Santiago de Cuba** (Z Escalona-Peña MD, M de los Ángeles-Ruban MD**,** N Sando-Antomarchi MD, M Dianela-Castillo MD**,** A Quintana-Polanco MD**,** L Ballester-Nolasco MD, I Cuevas-Torres BSc, L Borly-Parlay MD, K Corujo-Fong MD, A Díaz-León MD, M Leyva-Correoso MD, L Lopez-Gilbert MD, L Reyna-Bernal MD, Y Odio-Montero MD, GE Hechavarria-León MD, IM Bayard-Tellez MSc, O Borrero-Cobas MSc, M Biss-Savigne MSc, A Pico-López MSc, N Munder-Rubalcaba MSc, E Migue-Bonó BSc, I Sans-Labadi BSc, E Ferrer-Bicet BSc, Y Salabarria-Pérez BSc, L Mustelier-Correoso BSc, Y Poo-Santiesteban BSc, Y Roldan-Carvajal BSc, Y Echavarria-Mustelier MSc, Y Garbey-Rizo BSc, M Borrero-Palacio BSc, Y Ferrer-Vega BSc, A Hung-Colomar

Tech, T Ruiz-Iglesias BSc, K Hechavarría-Mena BSc, A Silva-Benavides BSc, Y Cruz-Fonseca BSc, G Zambrano-Betancourt BSc); **"Josué País" Polyclinic, Santiago de Cuba** (D Moraguez-Rodríguez MD, Y Silveira-Évora MD, I Ruiz-Guerrero MD**,** I Rizo-Sánchez MD, I Bermúdez-Revelo MD**,** M Donatién-Colón MD, O Camué-Lahera MD, D Valverde-Castillo MD, C Fernández-González MD, D Álvarez-Alba MD, Y Hervás-Ferrera MD, A I Despaigne-Bejerano MD, P I Garbey-González MD, C Rondón-Leyva MD, J E Álvarez-Tamayo MD, V Hechavarrìa-Michel BSc, T Caballero-González BSc, L Martínez-Garrido BSc, R Villavicencio-Guerrero MSc, N Rivaflechas-Sorrilla BSc, I Rojas-Mendoza BSc, O Odio-Pacheco BSc, I Gainza-Pajan BSc, K Hernández-La O BSc, A Jiménez-Franklin BSc, Y Barroso-Barroso BSc, N Massó-Villalón BSc, Y Carnota-Gomero BSc, V Jirón-Rivera BSc, N Bidet-Quesada BSc, S Machado-Jímenez MSc, M Verdecía-Benítez BSc, H Leyva-González BSc, M Preval-Cardona BSc, D García-Ricardo BSc, J Barroso-Duvergel Tech, Y Montero-Rodríguez Tech, Y Meriño-González Tech); **"Carlos J. Finlay" Polyclinic, Santiago de Cuba** (F Pasto-Rivero MD, Y Ricardo-Marzan MD, A B Romaguera-Perdomo MD, K Mustelier-Martínez MD, Y Vinent-Despaigne MD, EL Rey-Rovira MD, E Verdecía-Arocha MD, R Rodriguez-Carballoso MD, Z Torres-Rodríguez MD, G Matos-Rodríguez MD, G Ayala-Leal MD, B Carmenaty-Ferrer MD, M Uriarte-Vinent MD, M García-Ferrera BSc, Y Bello-Martínez BSc, C Labrada-Rivero BSc, Y Rodríguez-Milanés BSc, M Salazar-Reyes BSc, O Cabrera-Lago BSc, M I Ronaga-Aroche BSc, A Rodriguez-Lavigne BSc, I Fonden-Martínez BSc, B Estenoz-Odio BSc, Y Osorio-García BSc, N M Sancho-Monet BSc, E Y Pérez-Carrión BSc, M Mesa-Hechavarria BSc, Y M Heredia-Martínez MD, Y Meriño-Zayas MD, J Polanco-Rodríguez MD, Y Corría-Haber MD, Y Calzado-Cuesta Tech, E Montero-Martínez Tech); **"Camilo Torres" Polyclinic, Santiago de Cuba** (A L Cascaret-Santiago MD, I M Rosabal-Rosas MD, I Fresnedo-Canto MD, S D Somoza-Mograve MD, J F Rodríguez-Murillo MD, A Quintana-Batista MD, L González-Stevens MD, A León-González MD, E Ahuar-Gómez MD, C González-Aguilar MD, S Maturell-Hechavarria MD, E A Fonseca-Lambert MD, E R Carnota-Portela MD, K Benìtez-Nàpoles MD, L Acosta-Ochoa MD, A Dorado-Echavarría BSc, V Mourlot-Baubaire BSc, M E Silega-Romero BSc, Y Salazar-Denis BSc, M Bueno-Sierra BSc, Y Pozo-Rodríguez BSc, Y Pérez-Negret BSc, C M Menéndez-Román BSc, S Batista-Vázquez BSc, A Bosch-del Valle BSc, L Cedeño-Bles Tech, A Fuentes-Chávez MD, A García-Sarmiento MD); **"30 de noviembre" Polyclinic, Santiago de Cuba** (D Álvarez-Dupuy MD, I M Morejón-Rebelo MD, D Rodríguez-Mosqueda MD, Y Batista-Dales MD, N M León-Oran MD, M Guevara-Despaigne MD, C I Hechavarria-Mora MD, L Blanco-Cintra MD, R R Cleger-Bermúdez MD, S I Coloma-Oduardo MD, K Galán-Duany MD, D Almeida-Acosta MD, H M Guerrero-Delgado MD, S M Casin-Rodríguez MD, T Ramos-Acosta MD, A Hernández-Donatien MD, Y Alcolea-León MD, Y Savigne-Wilson MD, A M Álvarez-González MD, Y Valdez-Méndez MD, G González-Munded MD, A González-Galán BSc, Y Cuza-Molina BSc, E Meriño-Gutiérrez BSc, O A Cobo-Fonseca BSc, S E Ramos-Herrera BSc, E Vázquez-Castellanos BSc, Z Rondón-Cedeño BSc, M Carrión-Arias MSc, M Andrial-Mora MSc, M C Infante-Garzón MSc, R L Sarmiento-Hernández BSc, Y Medina-Caminero BSc, E Zapata-Blanco BSc, L Verguez-Ross BSc, Y Ramos-Lobaina BSc, K Faubel-Gómez BSc); **"Luis Ramírez López" Polyclinic, Santiago de Cuba** (T Céspedes-Baranda MD, D Cobo-Uriarte MD, I Cabrejas-Leguen MD, T Artílez-Uriarte MD, Y Calzado-Hernández MD, Y Zayas-Romero MD, L Puentes-Bordeloy MD, S Fong-Ortega MD, C Pérez-Borrero MD, M C Cruzata MD, LS Amiot-Gaskinz MD, M Lagar-Falcón MD, Y Rosabal-Sánchez MD, V Vera-Issac MD, Y Cintra-Vicente MD, N Ortega-Ojea MD, S Moreno-Velázquez MD, A Borrero-Nicot MD, Y A Hylton-Mena BSc, M Fernández-Maceo BSc, M Musle-Acosta BSc, A Claramunt-Alvarez BSc, A Mustelier-Montero Tech); **"Ernesto Guevara" Polyclinic, Santiago de Cuba** (R Montoya-Aranda MD, A Álvarez-Moraga MD, M Martínez-Luna MD, R A Rodríguez MD, L Aroche Maceo MD, D Díaz-Almenares MD, Y Torres-Hierrezuelo MD, Y Romero-Téllez BSc, P Ávila-Casamayor BSc, M Fuentes-Díaz BSc, O Santana-Gómez MSc, Y Vives-Acosta BSc, Y Ávila-Batista BSc, M Ross-Guevara BSc, S Aroche-La O BSc, J Aguilera-Perú BSc, I Verdazco-Almaguer BSc, E Rojas-Soulary BSc, A Lugo-Moret BSc, L D Abad-Pozo BSc, A Solari-Ocaña Eng, Y Toirat-Romani MD, A Riera-Ge MD, Y Fernández-Tamayo BSc, D Gual-Peña MD, B Pérez-Rivera MD, Y Giraudis-Kindelan MD, R Chaves-Toirat MD, R Reyes-Torres MD, L Villalón-Maturell MD, Y O Arzola-Vistel MD, D R Wilson-Aguilar MD, C Valverde-Ramón MD); **"Mario Muñoz" Polyclinic, Santiago de Cuba** (A Mesa-Yáñez MD, V Torres-López MD, A Sánchez-Cardero MD, I Savón-Rodríguez MD , G M Beltrán-Pérez MD, Y Montero-Montero MD, L Revilla-Díaz MD, L Torres-Quiñones MD, A J Hill-Correoso MD, D Castillo-Torres MD, J L Mustelier-González MD, S M Martínez-Gutiérrez MD, O Tamayo-García BSc, R Roldan-Rodríguez BSc, K Torres-Chaveco BSc, Y Lara-Zalazar BSc, O Álvarez-Rodríguez BSc, N Infante-Favier BSc, M E Meriño-Infante BSc, Y Castañeda-Oliva BSc, A G Despaigne-Hodelin BSc, G Sánchez-Lorchan BSc, V M Sánchez-Odren BSc, I López-Barroso BSc, L Bicet-Segura BSc, Y Brokart-Navarro MD, Y Tarrago-Hernández MD); **"Joaquín Castillo Duany" Hospital, Santiago de Cuba** (E Labrada-Diéguez MD, M Duconger-Danger MD, J A Castañeda-Fernández MD, R Pina-Núñez MD, A E Pérez-Cala MD, N Sánchez-Barrero MD, J Naranjo-López MD, G del Rio-Caballero PhD, M Suarez-Castañeda MD, S López-de Quesada MD, D Torres-Ramos MD, R E Despaigne-Salazar MD, E Call-Sayas MD, M Batista-Romagosa PhD, N Travieso-Ramos PhD, J C Maldonado-Avilez Eng, G Martínez-Arzola BSc, Y Fernández-Crespo BSc, D Zayas-Hechavarría BSc, Y Zamora-González BSc, Y Gómez-Coca BSc, N Mozo-Despaigne BSc, M C Reyes-Montero

BSc, D Núñez-Álvarez BSc, N Guzmán-Pérez PhD, N Sotelo-Sala BSc, Y Merbillet-Espinosa BSc); **"Asdrúbal López" Polyclinic, Guantánamo** (M Nordet-Torres MD, R Betancourt MD, Y C Cuesta-Millet MD, Y Columbié-Brooks MD, L Ramírez-López MD, R Heredia-Góngora MD, Y Creagh-Smith MD, D Pileta-Nápoles MD, S Machado-Realín MD, Y Armenteros-Carbonell MD, A Fuentes-Rodríguez MD, A Osoria-Sánchez MD, K Neyra-Garbey MD, D Rodríguez-Pérez MD, M Revé-Preval MD, B Betancourt-Abad MD, M Elias-Pichardo BSc, I Laffita-Gómez BSc, A Aguirre-Lambert BSc, Alicia Tito-Pérez BSc, M Matos-Ramírez BSc, Y Hernández-Martin BSc, D Tudela-Cascaret BSc, L Romero-Londres BSc, Y Lopez-Barrios BSc, I Conde-Casero BSc, M Rodríguez-García BSc, Y Hernandez-Peña BSc, A Favier-Elias BSc, A Rodríguez-Varga BSc, K Robas-López BSc, A García-Cedeño BSc, D Gómez-Romero BSc, J Samon-Pérez BSc, J Rodríguez-Guerra BSc, A López-Noa Tech); **"4 de abril" Polyclinic, Guantánamo (**A Reyes-Pacheco MD, W Brooks-Lestapier MD, A García-Fiol MD, A Leyva-Pérez MD, M Adams-Noblet MD, Y Illas-Bornot MD, J Sobrad-Vázquez MD, L Arroyo-Rodríguez MD, M Wilson-Labadí MD, J Domínguez-Figueroa MD, M Carcajal-Coello MD, R Paz-Bartelemi MD, E J Guilarte-Preval MD, E Palmero-Fuentes MD, M Álvarez-Rodríguez MD, L Serrano-Hardy BSc, M Palacios-Ortega BSc, J López-Megret BSc, Y Garcia-Alemán BSc, M González-Lobaina BSc, R Calderin-Sayut BSc, A Arrúe-Balón Tech, E Azahares-Leyva Tech, M Ramírez-Ramírez Tech, L Preval-Fernández Tech, Y Rodríguez-Columbié BSc, E Ramírez-Céspedes BSc, M Batista-Columbié BSc, Y Pérez-Ambruster BSc, L González-Brooks BSc, Y Laurencio-Cobas BSc, O Monterrey-Cejas BSc, R Gómez-Pérez Eng); **"Omar Ranedo" Polyclinic, Guantánamo** (A Fernández-Rodríguez MD, M Valiente-Guerra MD, R Matos-Díaz MD, J S Romero-Vargas MD, D Lao-Cárdenas MD, Y Elías-Oquendo BSc, Y Reyes-Romaguera MD, J Alba-Mosqueda MD, I Miraglia-Rosell MD, A C Durruty-Elías MD, A Soto-Franco MD, A Pérez-Leyva MD, K Sucet Elías-Armas MD, M Roche-Segura MD, I Manfarlane-Reyes MD, A E Olivares-Martínez MD, R González-Sierra BSc, N Cautin-Pineda BSc, M Rodríguez-Beltrán BSc, M Rojas-Rodríguez BSc, D Llamos-Leyva BSc, Y Lescaille-de la Cruz BSc, A Fasta-Estévez BSc, D Vinent-Portuondo BSc, Y Vallina-Fernández BSc, M Vega-Cuenca BSc, M de los A Paz-Camajuani BSc, Y Almenares-Súarez BSc, D Vidaillet-Rojas BSc, A Iglesias-Mizharrill MD); **"Mártires del 4 de agosto" Polyclinic, Guantánamo** (E Payam-Romero MD, M Vega-Fior MD, Y Bauza-Guerra MD, LR Rodríguez-Gallardo MD, Y Suarez-Alvarez MD, L Valera-Vigo BSc, M Quiala-Pérez MD, D Quintero-Castro MD, L Pérez-Calzado MD, A Leyva-Palacio MD, B Cosch-Duvergel MD, A V Hernández-García MD, A J Sánchez-García MD, C C Thompson-Acosta MD, D R Rodríguez-Zubizarreta MD, W Vázquez-Céspedes MD, R Johnson-Liben MD, F Castillo-Pérez BSc, D Leyva-Asin BSc, L Obregon-Sanchez BSc, L Sarmiento-Ramírez BSc, M Sánchez-Olivares BSc, S Ortiz-Ramírez BSc, O Ortiz-Fuentes BSc, D Issac-Argote BSc, N J Quintana-Reyes BSc, N Turro-Vigo BSc, M Sánchez-Castañeda BSc, E Speck-Speck BSc, A Barzaga-Vázquez BSc, M Raimon-Rodríguez BSc, E Ruiz-Batista BSc, N Pérez-Téllez Eng); **"Emilio Daudinot" Polyclinic, Guantánamo** (M Batista-Columbié MD, I Campos-Artiles MD, M Mallet-Verdecia MD, A de La Cruz-Despaigne MD, T Salazar-Quiñones MD, C C Sayoux-Thaureaux BSc, R F Planes-Ferrer MD, D C Omar-Borges MD, G J Cervera-Hernández MD, M E Girón-Morado MD, M Albelo-de la Cruz MD, A Savon-Ramírez MD, D Torres-Guerra BSc, V Fernández-Arnejo BSc, E Laffita-Pérez BSc, J Ceballo-Sánchez BSc, A Rodríguez-Noa BSc, I Estévez-Reyes BSc, T Cachimalle-Savon BSc, E Quiala-Ducas BSc, A Marsilli-Rivera BSc, Y Lau-Rodriguez BSc, W Díaz-Pérez BSc, R Alexander-Bodie BSc, M Veranes-Silven BSc, M Castillo-Prego BSc, D Duran-Áreas BSc); **"Jimmy Hirzel / Bayamo Oeste" Polyclinics, Granma** (R Marrón-González MD, I Castellano-Gómez MD, B C Addine-Ramírez MD, R A Rivero-Díaz MD, M E Parada-Escalona MD, C Guerra-Vázquez MSc, P Surós-Diaz MD, R B Fonseca-Muñoz MD, A Fonseca-Sanchez MD, C Hernandez-Torres MD, E V Romero-Fonseca MD, A D Vásquez-Machado MD, A M Millán-Guinarte MD, I Arévalo-Chavez MD, F R Rivero-Martinez MD, G Alvarez-Oliva MD, O Figueredo-Santos MD, M Olivares-Brito MD, M R Oduardo-Aguilar MD, D M García-Fernández MD, M Carrazana-López MD, Y Reyes-Méndez MD, Y Garcés-Hernández MD, R A García-González MD, G Reyna-Gomez MD, L C Pérez-Acevedo MD, A Corría-Barban BSc, D R Gibson-Guerra BSc, I Virelles-Espinosa BSc, A Pérez-Chávez BSc, D Gibson-Guerra BSc, MC García-Blanco BSc, N R Varona-Amaya BSc, J Fonseca-Santos BSc, J Vargas-Monpie BSc, A Cedeño-Reyes BSc, Y Hernández-Pérez BSc, O Guzmán-Montero BSc, S Núñez-Rodríguez BSc, M C Quiala-Hechavarria BSc, M M Vega-Blanco BSc, D Almenares-Zamora BSc, M Rosabal-Antúnez BSc, X Alvarez-Martínez BSc, G C Zamora-Figueredo BSc, Y Rodríguez-Díaz BSc, R L Moya-Calisté, BSc, Y Fabre-Gil BSc, E Serrano-Rodríguez BSc, Y González-Fonseca BSc, S Díaz-Cruz BSc, E Cervantes-Rondón BSc, M E Tamayo-Espinosa BSc, C Ramos-Ayala BSc, A M Fonseca-Fleitas Tech, A Rodríguez-Pinos BSc, D Y Machado-Zamora BSc, Y Fonseca-Rivero BSc, C Tamayo-Zamora BSc, V D Rodriguez-Morales BSc, D Peña-Correa BSc, A M Zaldívar-Suárez BSc, M A Licea-Barban BSc, Y Barroso-Solano BSc, Y Fidalgo-Domínguez BSc, A Gardón-Expósito BSc, D L Pompa-Ramírez BSc, Y Vinajera-Milán BSc, V Viamonte-Piña BSc, R M Enamorado-Saldaña Tech, I C Gonzalez-Olive Tech, E Fonseca-Milanés BSc, A I Espinosa-Guerra BSc); **"Arnaldo Milián" Hospital, Santa Clara, Villa Clara** (J G Martínez-Urbay MD); **"Mártires del 9 de abril" Hospital, Sagüa la Grande, Villa Clara** (N L Vasallo-Hernández MD); **National Coordinating Center for Clinical Trials, Santiago de Cuba** (D Y Griñan-Semaná MSc, A Arteaga-García MD); **Provincial Health Directorate, Santiago de Cuba** (M del Carmen Calzado-Alcolea

MD, A Hernández-Magdareaga MD, A Legrá-Fernández Eng); UCT Geocuba Investigación y Consultoría (P M Velazco-Villares MSc); **Centre for Genetic Engineering and Biotechnology, Havana, Cuba** (E García-Iglesias BSc, A Álvarez-Acosta MSc, Y Duncan-Roberts MD, C Martínez-Suárez BSc, O González-Díaz MD, V Frómeta-Alberti BSc, D Rojas-Socorro Tech, Z Santana-Vázquez MSc, C Serrallonga-Trujillo BSc, C Chuay-Silva Eng, Y Crespo-García BSc, U Garriga-Pereira Eng, G E Guillén-Nieto PhD, R Ricardo-Parellada MSc, D Rodríguez-Reinoso M Calderín-Ricardo Tech, T Díaz-Argudín Tech, M Ale-Martínez Tech, L C Bakos-Ruíz BSc, Y Delgado-Piedra BSc, G Lemos-Pérez MSc, J L Vega-Elías Eng); **Drug Marketing and Distribution Company – EMCOMED, Cuba** (A L Chacón-Padrón BSc, Y González-Olivera BSc); **AICA Laboratories Company** (A E Vallin-García MSc); **Immunoassay Center, Havana, Cuba** (A Palenzuela-Díaz MSc, I Y Valdivia-Alvarez PhD); **Virology Laboratory, Provincial Centre for Hygiene, Epidemiology and Microbiology, Santiago de Cuba** (M J Gadillo-Pérez MSc, R Zalazar-Quevedo BSc, D Benitez-Fernández BSc, L Delgado-González MSc, Y Mederos-Nuñez BSc, M J Rodríguez-Díaz BSc, D L García BSc, J Palacios-Silveira BSc, P L Hechavarría-Galán MD, M Cobas-Soler Tech, M V Soler Pérez BSc); **Institute of Cybernetics, Mathematics and Physics, Havana, Cuba** (Prof J E Sánchez-García PhD, R Selgas Lizano BSc, J L Azor Hernández MSc); **Biotechnology and Pharmaceutical Industries Group, BioCubaFarma, Havana, Cuba** (E Martínez-Díaz PhD).
